# Covid-19 SEIDRD Modelling for Pakistan with implementation of seasonality, healthcare capacity and behavioral risk reduction

**DOI:** 10.1101/2020.09.01.20182642

**Authors:** Shoab Saadat, Salman Mansoor

## Abstract

**INTRODUCTION:** *Introduction:* December 2019 saw the origins of a new Pandemic which would soon spread to the farthest places of the planet. Several efforts of modelling of the geo-temporal transmissibility of the virus have been undertaken, but none describes the incorporation of effect of seasonality, contact density, primary care and ICU bed capacity and behavioral risk reduction measures such as lockdowns into the simulation modeling for Pakistan. We use above variables to create a close to real data curve function for the active cases of covid-19 in Pakistan.

*Objective:* The objective of this study was to create a new computational epidemiological model for Pakistan by implementing symptomatology, healthcare capacity and behavioral risk reduction mathematically to predict of Covid-19 case trends and effects of changes in community characteristics and policy measures.

*Methods:* We used a modified version of SEIR model called SEIDRD (Susceptible - Exposed Latent - Diagnosed as Mild or severe - Recovered - Deaths). This was developed using Vensim PLE software version 8.0. This model also incorporated the seasonal and capacity variables for Pakistan and was adjusted for behavioral risk reduction measures such as lockdowns.

*Results:* The SEIDRD model was able to closely replicate the active covid-19 cases curve function for Pakistan until now. It was able to show that given current trends, though the number of active cases are dropping, if the smart lockdown measures were to end, the cases are expected to show a rise from 28th August 2020 onwards reaching a second peak around 28th September 2020. It was also seen that increasing the ICU bed capacity in Pakistan from 4000 to 40000 will not make a significant difference in active case number. Another simulation for a vaccination schedule of 100000 vaccines per day was created which showed a decrease in covid cases in a slow manner over a period of months rather than days.

*Conclusion:* This study attempts to successfully model the active covid-19 cases curve function of Pakistan and mathematically models the effect of seasonality, contact density, ICU bed availability and Lockdown measures. We were able to show the effectiveness of smart lockdowns and were also to predict that in case of no smart lockdowns, Pakistan can see a rise in active case number starting from 28th of August 2020.

## 1. Introduction

### 1.1 Background

December 2019 saw the origins of a new Pandemic which would soon spread to the farthest places of the planet. Several efforts of modelling of the geo-temporal transmissibility of the virus have been undertaken, but none describes the incorporation of effect of seasonality, contact density, primary care and ICU bed capacity and behavioral risk reduction measures such as lockdowns into the simulation modeling for Pakistan. The aim of this study was to create a new computational epidemiological model for Pakistan by implementing symptomatology, healthcare capacity and behavioral risk reduction mathematically to predict of Covid-19 case trends and effects of changes in community characteristics and policy measures. To achieve this we propose a novel SIR-type metapopulation transmission model and a set of analytically derived model parameters.

### 1.2 Objective

The objective of this study was to create a new computational epidemiological model by implementing symptomatology, healthcare capacity and behavioral risk reduction mathematically to predict of Covid-19 case trends and effects of changes in community characteristics and policy measures.

## 2. Methods

### 2.1 Data Resources

Real world COVID-19 Data utilized for this study can be found on the following repository maintained by the Center for Systems Science and Engineering (CSSE) at Johns Hopkins University and “Our World in Data” website page “Coronavirus Pandemic (COVID-19)” https://github.com/CSSEGISandData/COVID-19 https://ourworldindata.org/coronavirus

### 2.2 Model Compartmentalization

We used a modified SEIR model with slight modifications in the compartmentalization and parameterization of an existing example (1). We call this model SEIDRD for the presence of susceptible, exposed-latent, infectious, detected-symptomatics (mild or severe), recovery and deaths compartments (Table 2). Following is detailed look into these compartments:

**Table 1.**
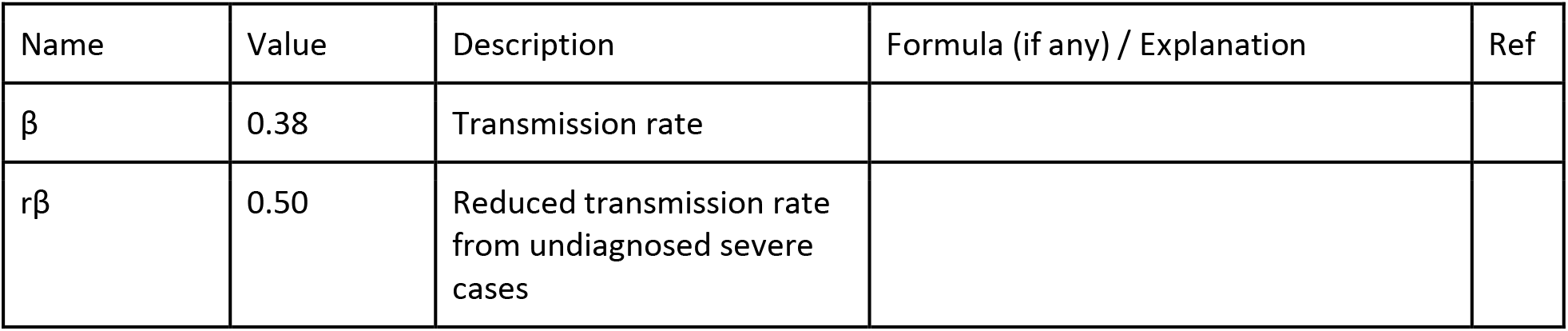
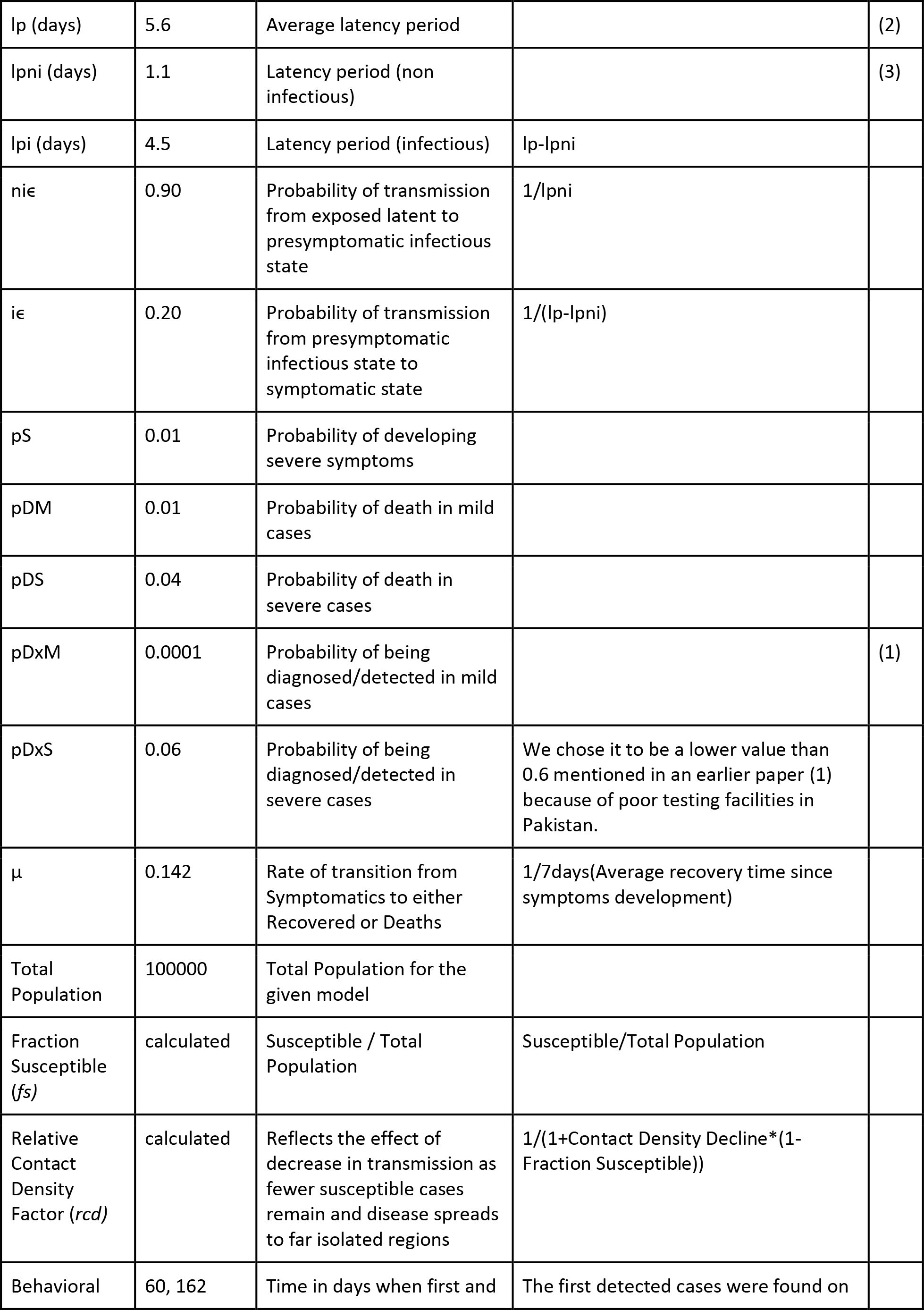
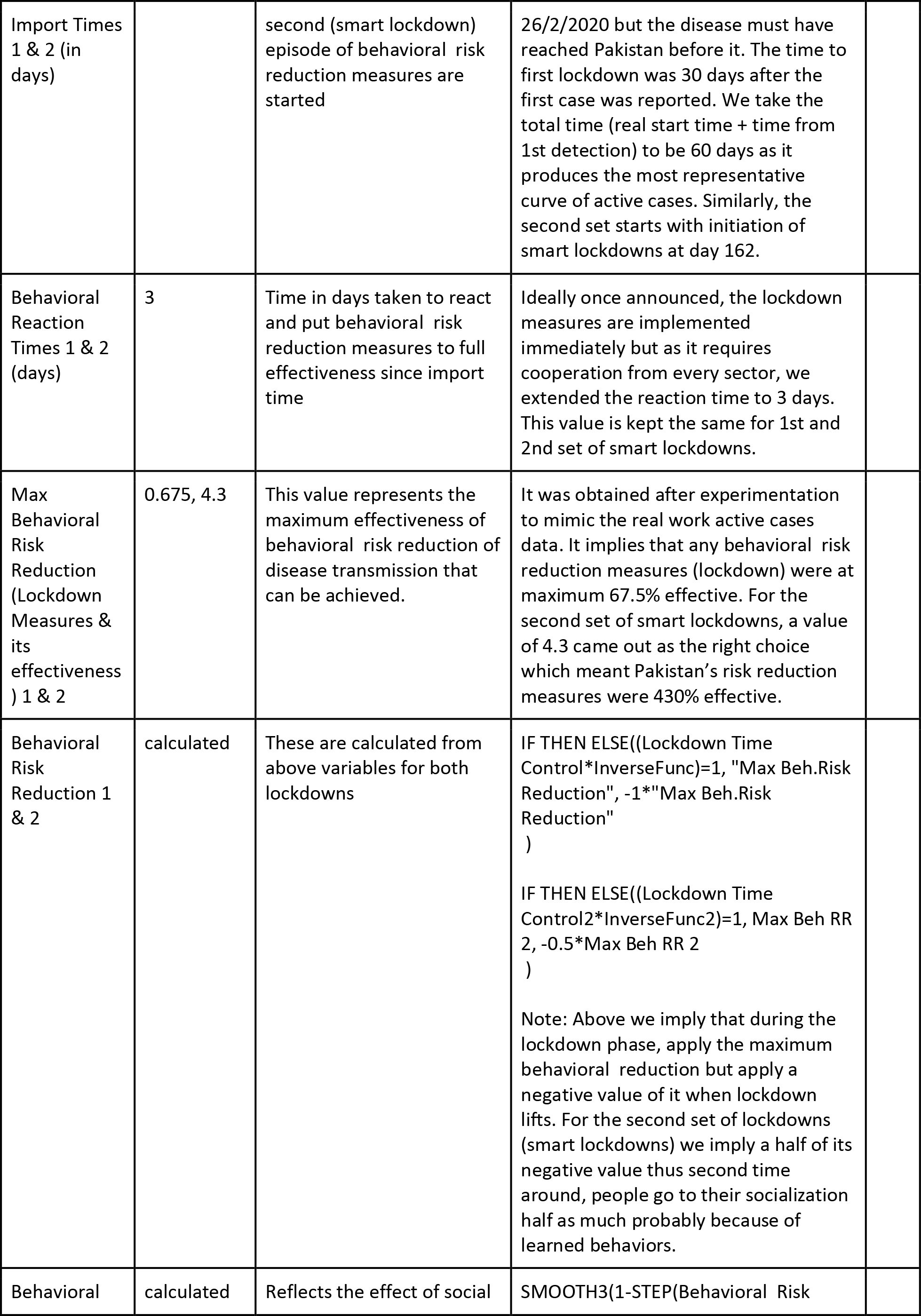
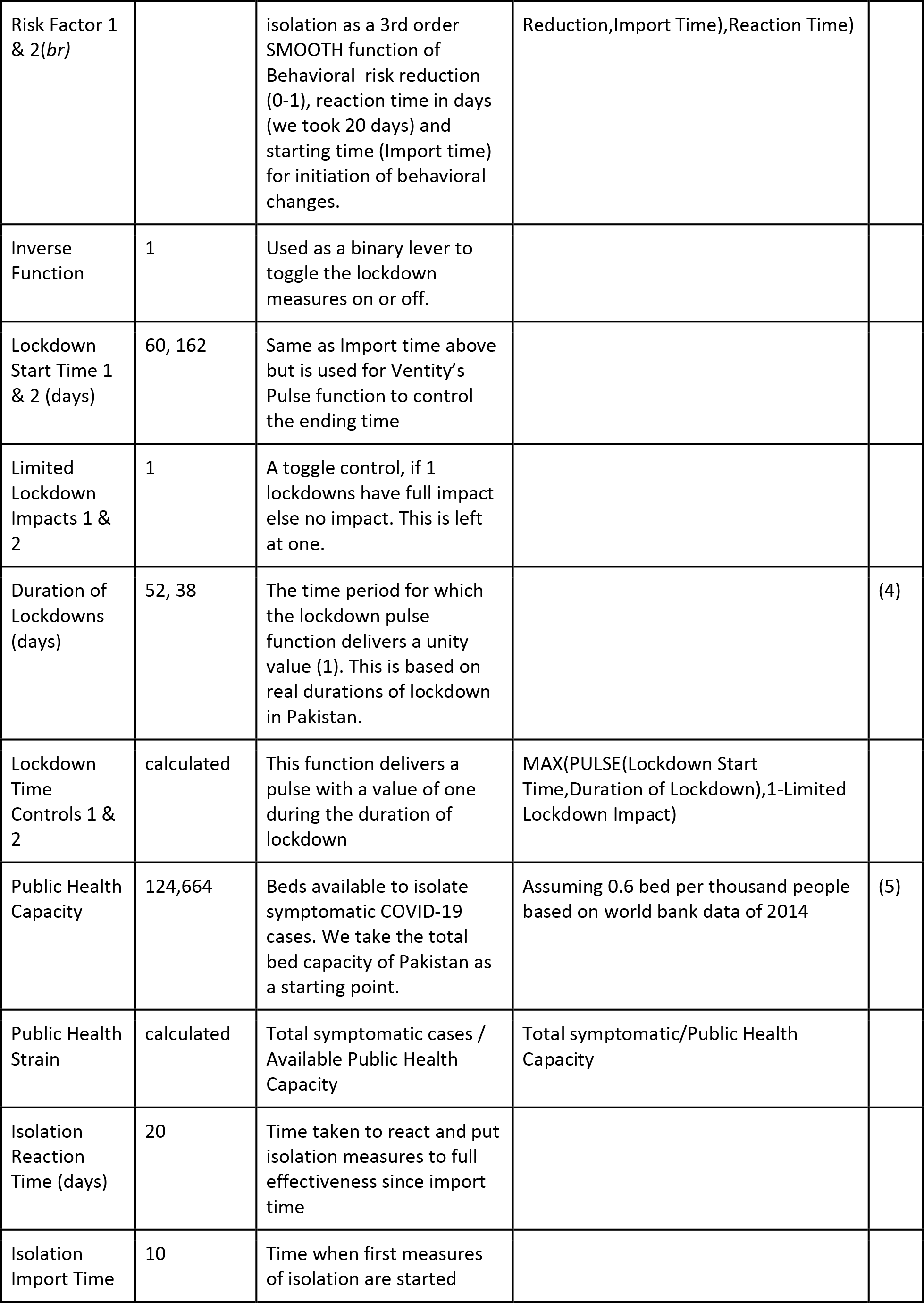
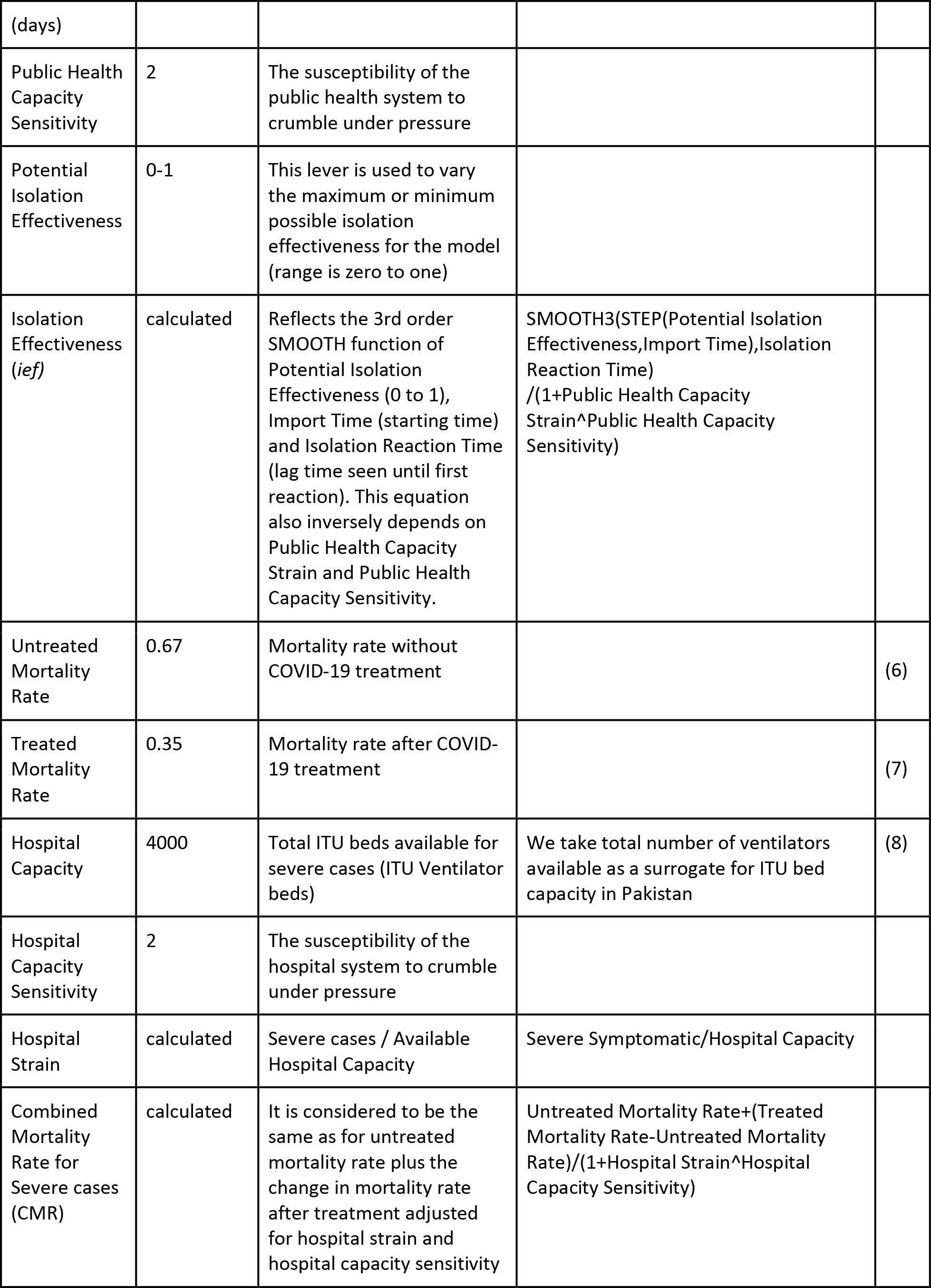
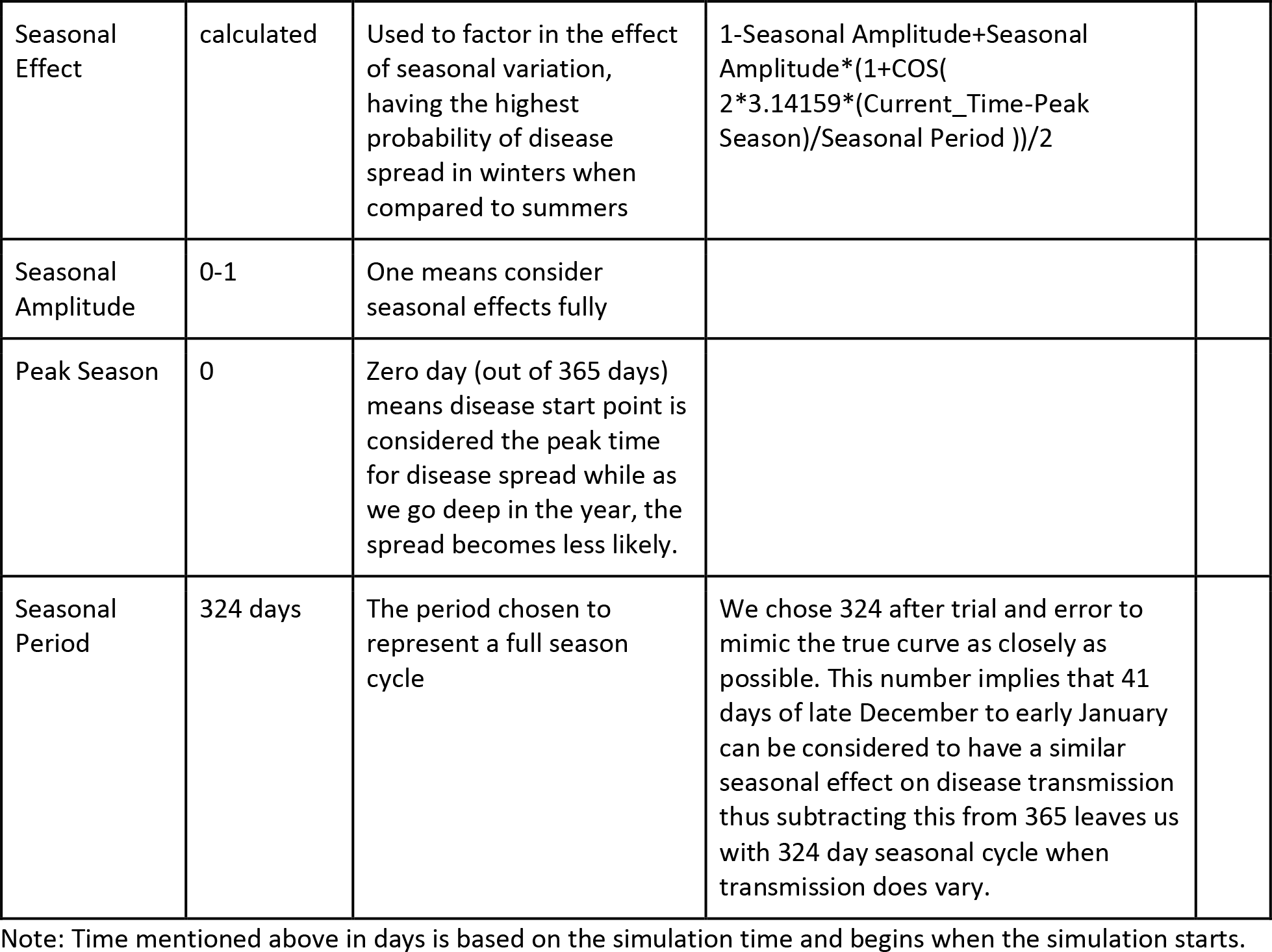
Parameter values used

**Table 2.** Compartments and transition equations used. Note: (_t) is used for all transition equations at the end of symbol names

| Name | Formula | Description | Ref |
| --- | --- | --- | --- |
| Total Population (tP) | 207,774,520 | Starting population chosen to be Pakistan's population. |  |
| Susceptible (S) | $S - E_t$ | Total susceptible population | |
| Exposing ($E_t$) | $\frac{\partial S}{\partial t} = (fs.br.rcd). (PI(\beta) + MS(\beta)(1 - ief) + SS(r\beta)(1 - ief))$ | Change in susceptible population which are transitioning into exposed population over the given time period $t$ | |
| Exposed-Latent (E) | $E_t - I_t$ | Patients exposed to Covid-19 but have neither yet developed the symptoms nor can they spread the disease. | |
| Infecting ($I_t$) | $\frac{\partial E}{\partial t} = E.nie$ | Change in exposed populations which are transitioning into the presymptomatic infectious population. | |
| Presymptomatic Infectious (PI) | $I_t - MC_t - SC_t$ | Patients who can spread the disease but haven't yet developed the symptoms. | |
| Mild Converting (MC<sub>t</sub>) | $\partial PI(MC_t)/\partial t = PI \times i_e \times (1 - pS)$ | Transitioning population from presymptomatic infectious to Mild Symptomatics | |
| Severe Converting (SC<sub>t</sub>) | $\partial PI(SC_t)/\partial t = PI \times i_e \times pS$ | Transitioning population from presymptomatic infectious to Severe Symptomatics | |
| Total converting (TC<sub>t</sub>) | $\partial PI/\partial t = MC_t + SC_t$ | The total of Mild and Severe symptomatic transitioning populations | |
| Mild Symptomatic (MS) | $MC_t - MR_t - MD_t$ | Patients with mild symptoms of the disease | |
| Severe Symptomatic (SS) | $SC_t - SR_t - SD_t$ | Patients with the severe symptoms of the disease | |
| Total Symptomatic (TS) | $MS + SS$ | Sum of mild and severe symptomatic populations | |
| Milds Recovering (MR<sub>t</sub>) | $\partial MS(MR)/\partial t = MS \times (1 - pDM)$ | Transitioning population from Mild Symptomatic population to the Mild Recovering population | |
| Milds Dying (MD<sub>t</sub>) | $\partial MS(MD)/\partial t = MS \times pDM$ | Transitioning population from Mild Symptomatic population to the Mild Dying population | |
| Severe Recovering (SR<sub>t</sub>) | $\partial SS(SR)/\partial t = SS \times (1 - pDS)$ | Transitioning population from Severe Symptomatic to Severe Recovering population | |
| Severes Dying (SD<sub>t</sub>) | $\partial SS(SD)/\partial t = SS \times pDS$ | Transitioning population from Severe Symptomatic to Severe Dying population | |
| Recovered (R) | $MR_t + SR_t$ | Recovered cases | |
| Deaths (D) | $MD_t + SD_t$ | Total deaths | |

**Figure 1.**
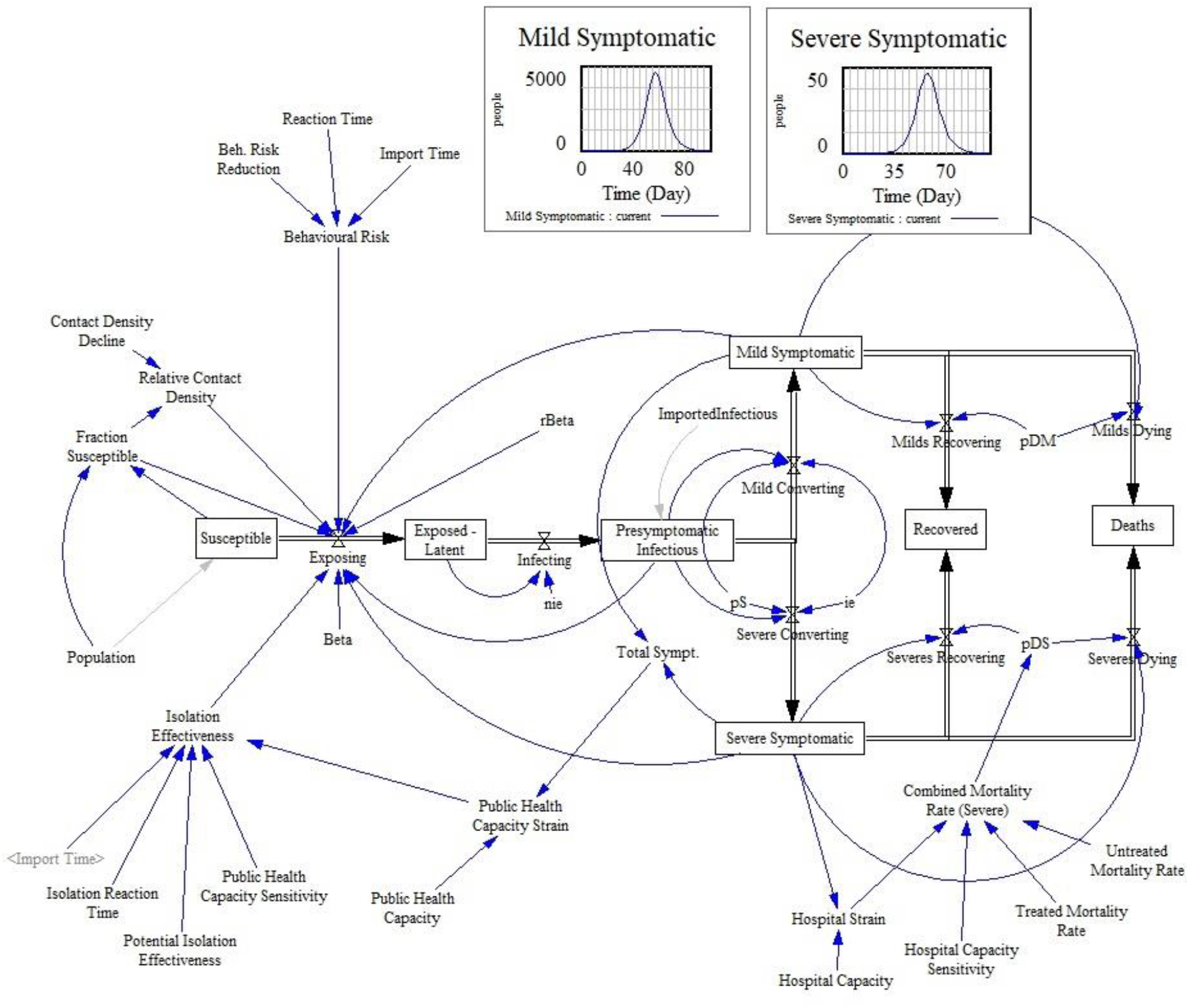
Basic Compartmental modeling setup with SEIDRD compartments arranged as above.

#### 1. Susceptible (S)

This represents the population set which is susceptible to COVID-19. Any exposure to Presymptomatic Infectious (PI), Mild Symptomatic (MS) or Severe Symptomatic (SS) populations can lead to a conversion of susceptible persons to an Exposed-Latent (E) patient with a probability of Beta (β) (Table 1). The transition equation for this population’s conversion into Exposed-Latent population is given as:

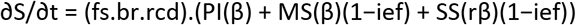

In this equation, fs represents Fraction susceptible, br is behavioral risk factor while rcd represents relative contact density (Table 1). Each of these parameters are additional controlling levers which can either increase or decrease the rate of conversion into Exposed-Latent population. A decrease in fs reflects less susceptibility to infection, low br reflects better hygienic and social isolation behaviour while a low rcd reflects sparse population density when number of susceptibles decreases, thus naturally low contact between the remaining susceptibles as they are far apart. Decrease in any or all of these parameters can slow down the rate of conversion into the next compartment. The second part of the equation represents the ability of PI, MS and SS to infect a susceptible person with the probabilities of β, (β)(1-ief) and (rβ)(1-ief) (Table 1) respectively. β reflects the probability of getting infected from a PI population, rβ represents the relative change in probability of getting infected if the infection is coming from a severe case to a susceptible, while (1-ief) is a measure of isolation effectiveness (Table 1). Isolation effectiveness (ief) is a measure of effectiveness of isolating total symptomatic patients. It is one if every symptomatic patient is completely isolated and cannot infect any other susceptible person. It does not cover the PI population as they are usually not identified unless symptomatic thus does not have a huge impact.

#### 2. Exposed-Latent (E)

This population represents the patients who have acquired the disease but are neither showing any symptoms nor able to transmit the disease. This population converts into Presymptomatic Infectious population. The transition equation is given as:

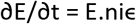

Here, niϵ represents non-infective-epsilon which represents the probability of transmission from exposed latent to presymptomatic infectious state.

#### 3. Presymptomatic Infectious (PI)

These are the patient population who have acquired the disease and are infectious to others but have not developed any symptoms yet. Thus, they usually remain under the healthcare radar and are usually the most common source of disease spread. They transition into either Mild Symptomatic cases of Severe Symptomatic ones. The transition equations is as follows:

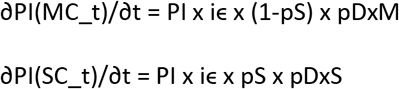

Here δPI(MC_t)/δt represents the rate of change of PI into Mild Symptomatic cases while δPI(SC_t)/δt represents the transition into Severe symptomatic cases. The probability of conversion from PI into the next compartment is represented by iϵ (infective-epsilon) while out of this converted population, the probability of converting into Severe Symptomatic population is given by pS. The probability of detection / diagnosis is given by pDxM and pDxS for mild and severe cases respectively (Table 1).

#### 4a. Mild Symptomatic (MS)

This compartment represents the patients who have now developed mild symptoms and are most likely to either recover or a minority could still die. Being mildly symptomatic, they can also infect the susceptible population with a rate of (β)(1-ief) if isolation effectiveness is not one as described above. The transition equations for this compartment are as follows:

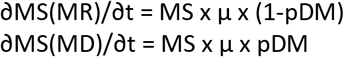

Here ∂MS(MR)/∂t represents the rate of change of MS cases into Recovered population with a total probability of μ x 1-pDM while ∂MS(MD)/∂t represents the conversion of MS cases into the cases who die with a total probability of μ x pDM (Table 1). Mu (μ) represents the probability of transitioning from any symptomatic case into any next compartment.

#### 4b. Severe Symptomatic (SS)

This represents the severe cases compartment which are converted from PI compartment. This compartment then transitions into either recovered cases or deaths as given by following equations respectively:

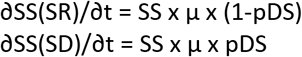

Here, ∂SS(SR)/∂t represents the rate of conversion of SS population into recovered ones with a probability of (1-pDS) while ∂SS(SD)/∂t represents the rate of conversion of SS population into deaths with a probability of pDS (table 1). The probability of deaths in severe cases (pDS) is in turn calculated as combined mortality rate (CMR) (Table 1). This CMR variable is in effect determined by minimal rate of mortality (as in mild cases) plus any additional mortality risk imparted by the lack of hospital capacity. Thus, if hospital strain (Severe cases / Available Hospital Capacity) is high, there is additional mortality risk imparted and thus the mortality in severe cases will be at-least the same as in treated mortality plus an additional risk. Its maximum value can get as high as the untreated mortality (Table 1).

#### 5. Recovered (R)

This compartment represents all those who have recovered after being mildly or severely symptomatic.

#### 6. Deaths (D)

This represents total deaths observed as converted from either mild or severely symptomatic compartments.

**Figure 2.**
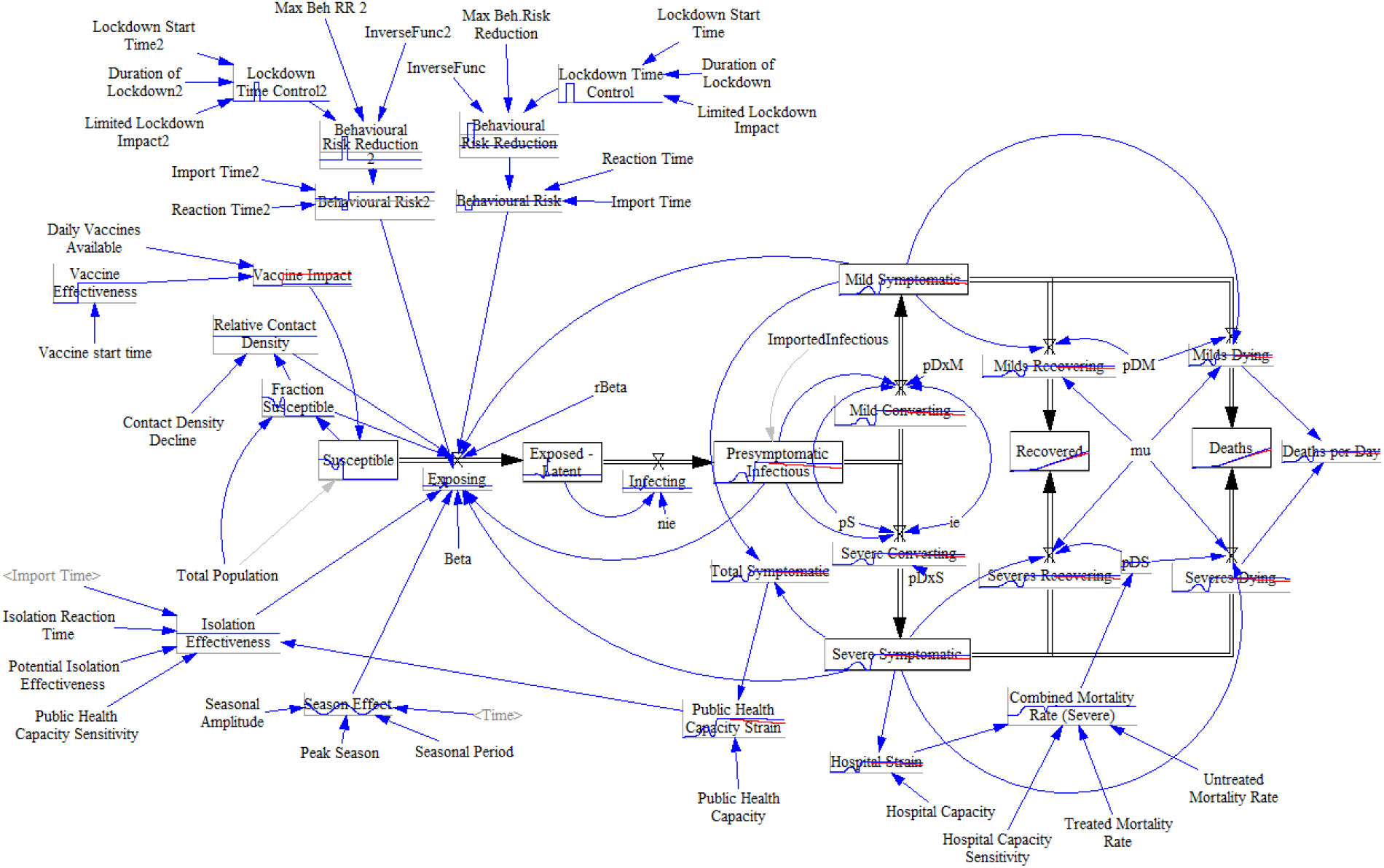
Full SEIDRD Modeling chain. It shows the implementation of Hospital strain, Public health capacity strain, Seasonal effect, Isolation effectiveness, Relative contact density, Vaccine effectiveness and Lockdown 1 & 2 as important variables affecting the final prediction.

### 2.3 Model Parameterization

Above in Table 1, we present the parameters chosen for the model. These have been derived after literature review and represent either the best estimates mentioned in the literature, or most plausible values by the authors based on the epidemiological knowledge on SARS-CoV-2 and other viruses.

#### Literature based evident parameters

We split the total time between a person gets infected and develops symptoms into non infective latent period (ni) and infective (i) latent periods. We then calculate the parameters nie and ie, using the estimates of average latency period (lp) (2) and non infective latency period (lpni) (Wallinga & Teunis, 2004) as given by the formulae in Table 1. We chose to use Korean proportion of “severe” to diagnose cases as a base for the probability of developing the severe condition (pS), and we set it to 0.01.

Among other important parameters were β, rβ and μ which represent the effective contact rate, reduction in contact rate in severe cases and rate of transitioning from symptomatic cases. β can be calculated from previous parameters as:

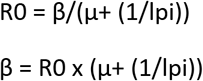

There are widely varying estimates for R0 in literature with values ranging from 1.4 to 6.49 (10-14) We decided to choose the R0 of 4.4 reflecting a relatively higher rate of spread which is well within the range of 2-5, modelled 223 for SARS (3). We derive μ from a safe quarantine period for diagnosed cases equal to 10 days (15). We assumed 1/μ to last on average for 7 days from symptoms development to recovery. The sum of1/ μ and previously estimated lpi (presymptomatic infectious period) results in 11.5 days which represents the total duration from becoming infectious till recovery after a person acquires the disease. Using μ and (1/lpi), we can now calculate β which comes equal to 0.383. We also selected rβ to be 0.5 following the assumption for this parameter used in the 2009 influenza outbreak (16). This is because patients who have severe disease are mostly admitted, thus isolated and have reduced rates of transmission.

The probability of death or mortality rate varies widely from values around 0.01 to 0.1 based on age and other parameters (17), (18), (19). For rate of mortality in mild cases (pDM), We chose a value on the lower spectrum of 0.01, based on the results obtained from early Wuhan studies (20). For severe cases admitted in ITUs, the mortality rate spectrum ranges from 0.11 to 0.42 depending on the age group with an average of 0.35 (21). In one study, the mortality rate in ICU patients was reported as high as 0.67 (6). In order to calculate the combined rate of mortality in severe cases (CMR) (Table 1), we used two types of mortality estimates: 1. Untreated mortality rate, which was chosen to be 0.67 being on the higher end of the spectrum, 2. Treated mortality rate, which was set at 0.35, assuming it to be on the lower end of the spectrum but still representing an average value.

### Local conditions based estimated parameters

In order to analyse the model for Pakistani population we choose the parameters based on the most recent available literature as given in Table 1 with references. We choose the total population to be 207,774,520. Based on a report from the World bank, the total bed capacity for Pakistan was calculated to be 0.6 per thousand (5). We chose this value to represent the total public health capacity as in the ideal case scenario, any symptomatic case will be admitted and isolated for the disease duration. For the purpose of hospital capacity variable, we selected the total number of ventilators (4000) as a surrogate for the total ICU beds in Pakistan (8).

**Figure 3.**
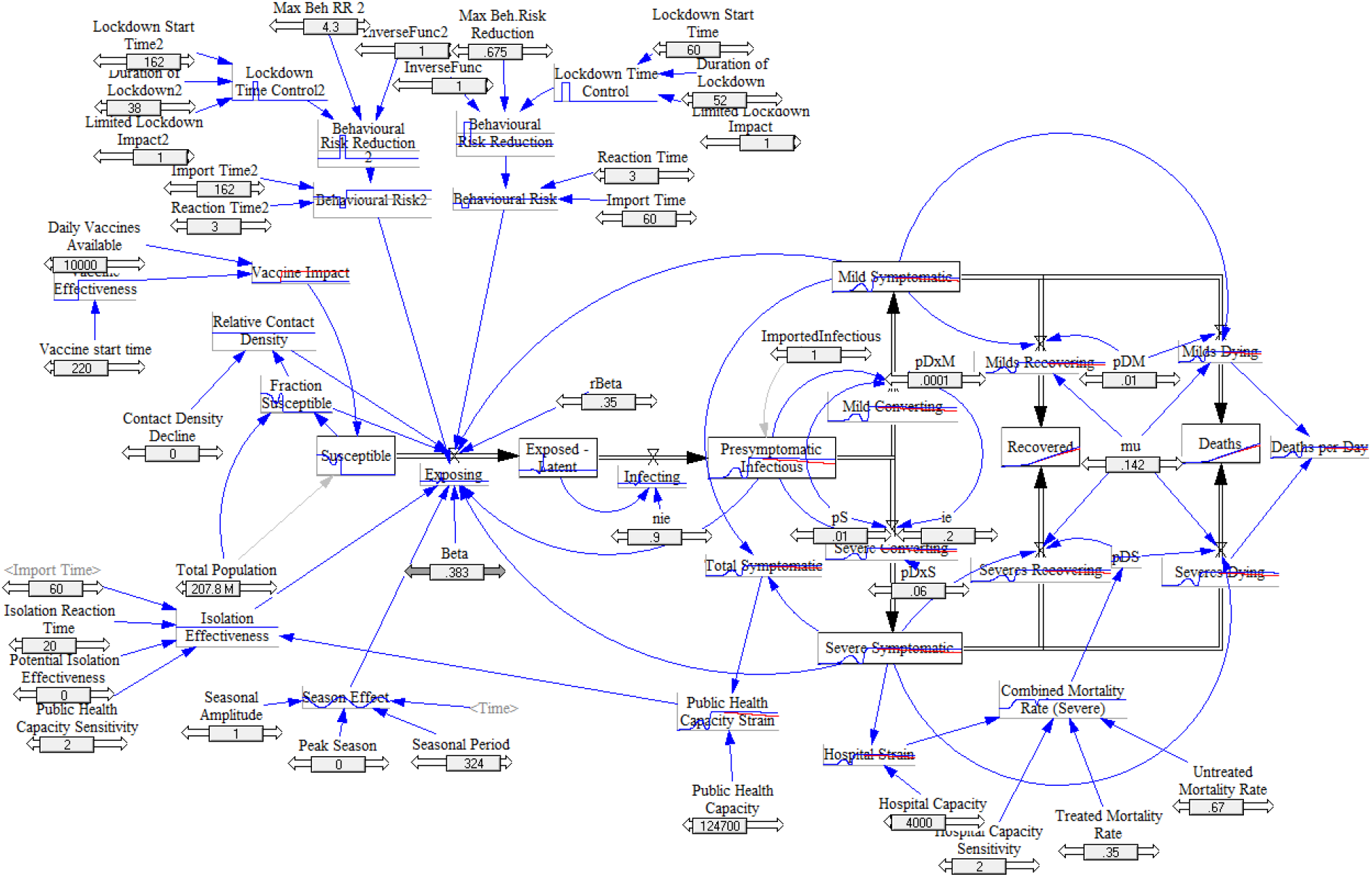
Detailed SEIDRD Model with default initialization values.

### 2.4 Modelling with Vensim PLE (version 8.0)

We use Vensim PLE software version 8.0 (http://www.vensim.com) for the model development and numerical integration. Vensim, the Ventana Simulation Environment, is an integrated framework for conceptualizing, building, simulating, analyzing, optimizing and deploying models of complex dynamic systems (22) For our purpose, we use first order Euler integration technique to solve the equations numerically which is built into the software (23). The simulation period was selected to be 12 months.

## 3. Results & Discussion

In order to simulate the Covid-19 curve of Pakistan, some of the assumptions in Table 3 were made. We then used the constants and variables as defined in Table 1 and 2 to come up with the most similar curve of total symptomatic (reported) cases in Pakistan. We divide the derivation into following experimental stages:

**Table 3:** Assumptions Made

| Compartment | Assumption |
| --- | --- |
| Total Population | Whole population exists in a homogeneous area without any provincial or regional compartmentalization |
| Susceptible | No existing immunity to infection. |
| Mild Symptomatics | For model simplicity, we decided to merge into one compartment all mild and asymptomatic cases.<br>Patients with mild symptoms, in contrast to those in severe condition, are still |
|  | 172 capable of travelling. |
| Behavioral Risk Reductions / Lockdowns | After experimenting with the values, we assumed that during lockdown phases, people have a reduction in spread of active cases by a factor of Behavioral risk reduction (BRR) value but non lockdown periods, people often go back to socialization and spread of disease activity is increased by the BRR value. |
| Time to react | We assume that it takes around 3 days for people to properly respond to any risk reduction measures like lockdowns |
| Hospital Capacity | We assume that beds needed for severe cases equates to ITU beds available. |
|  | Everyone who gets infected is removed from the population either through recovery or death. |
|  | The population is large, fixed in size and it is confined geographically. |
| Recovered | Recovered individuals cannot be reinfected, although the only evidence so far is for rhesus macaques (Ota, 2020) and WHO is still investigating the issue. |

Experiment 1 creates a base model with seasonal variation incorporated and shows the expected curve if none of the lockdown measures or risk reduction methods were implemented. In experiment 2, we implement the first set of lockdown measures and see their effect without the subsequent smart lockdowns.

In the third experiment, we fully implement the smart lock downs as well and observe the similarity between actual case number and our predicted ones. In the fourth experiment, we test two assumptions; 1. What if the smart lockdown measures end? 2. What if some form of smart lockdown measures persist. Finally, In the fifth experiment, assuming that covid-19 returns, we observe the effect of varying hospital, public health capacity and possible vaccination and its impact. Before reviewing the following experiments, it’s important to note that the simulation time starts 36 days before the first 2 cases were officially reported in Pakistan (Table 4), thus any of the day references mentioned below are from the simulation timescale.

**Table 4.** Pakistan Initiatives and disease response timeline

| Date | Simulation equivalent time | Initiative | Actual Cases | Active Infectious (Total symptomatic diagnosed) cases as per our model | Ref |
| --- | --- | --- | --- | --- | --- |
| 26/2/2020 | 36 | First Reported Cases | 2 | 2 | (9) |
| 20/3/2020 | 59 | First Reported Death | 485 | 346 |  |
| 24/3/2020 | 63 | First Lockdown starts in Capital | 947 | 775 |  |
| 26/3/2020 | 65 | Cases Reach 1000+ | 1171 | 1146 |  |
| 26/4/2020 | 96 | Cases Reach 10000+ | 10111 | 11201 |  |
| 9/5/2020 | 109 | Lockdown Ends | 20290 | 18655 |  |
| 9/6/2020 | 140 | Cases Reach 70000+ | 71127 | 75260 |  |
| 19/6/2020 | 150 | Cases Reach 100000+ | 100450 | 92916 |  |
| 27/6/2020 | 158 | Starts to plateau | 107942 | 99954 |  |
| 1/7/2020 | 162 | Highest case number & Smart Lockdown starts | 108273 | 101668 |  |
| 3/7/2020 | 164 | Cases start decreasing | 103722 | 102212 |  |
| 10/8/2020 | 202 | Smart lockdown ends | 17799 | 22264 |  |

Note: The root mean squared error for above table equals 4157. This is high for the early phase of the disease but relatively small when total cases get to a hundred thousand mark.

**Figure 4.**
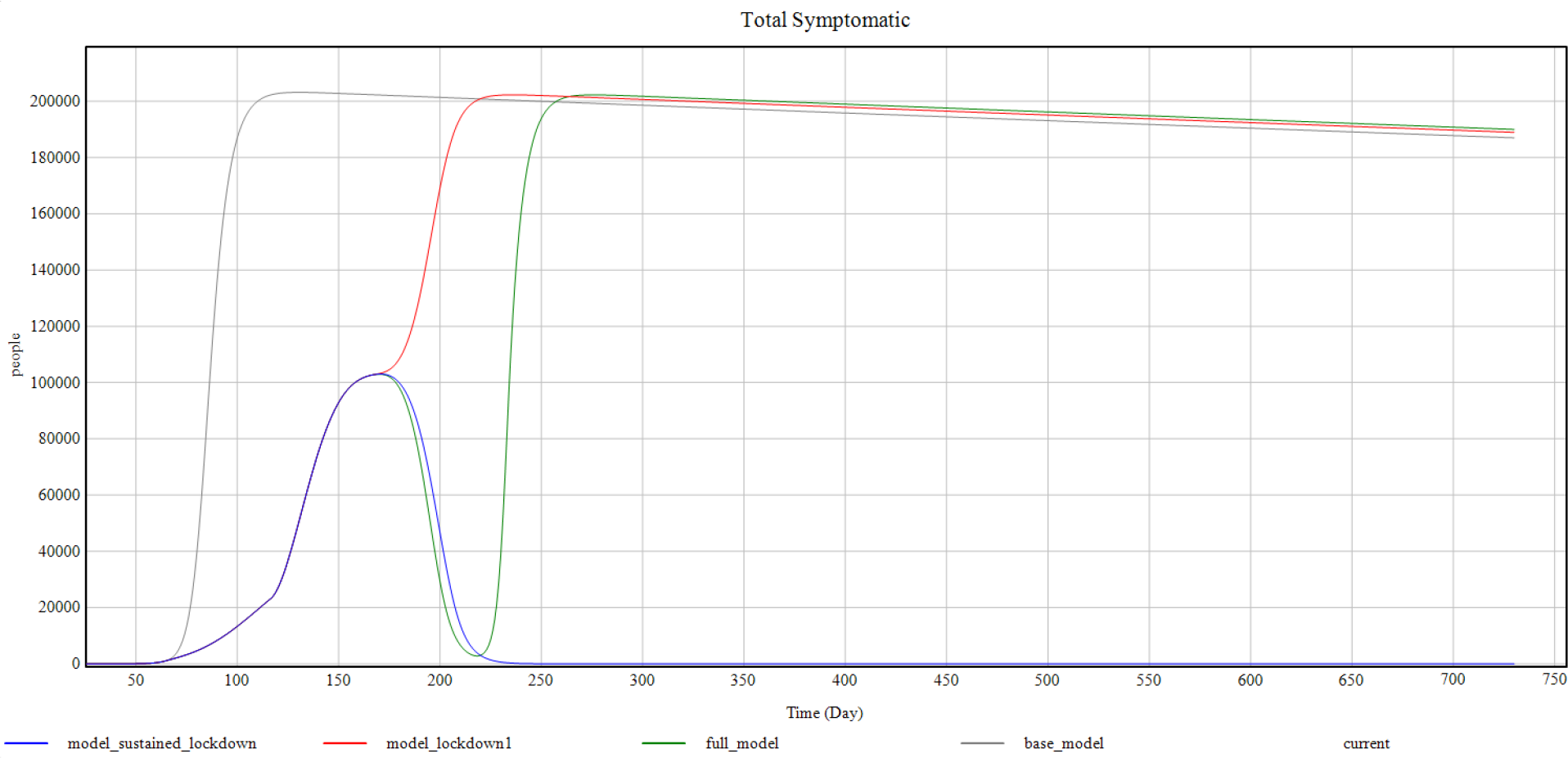
Shows the combined prediction curves for total symptomatic cases against simulation time. It compares base model, experiment 2, 3 and 4 models side by side.

### Experiment 1 (Base Model - No Lockdowns)

In this experiment, we run the simple base model consisting of SEIDRD compartments as explained in the methodology section. This run was under the assumption that none of the lockdown measures have been imposed and the cases are spreading at a natural rate. This model also incorporates the seasonality adjustment (Table 2). As per this model, the cases start rising around day 70 to 75 simulation time. The cases peak at around day 125 with a maximum of around 200000 cases being reported each day. The numbers then start to decline but very slowly.

As this model assumes no behavioral risk reduction measures, this would have been the worse case scenario were it not for the timely lock downs.

**Figure 5.**
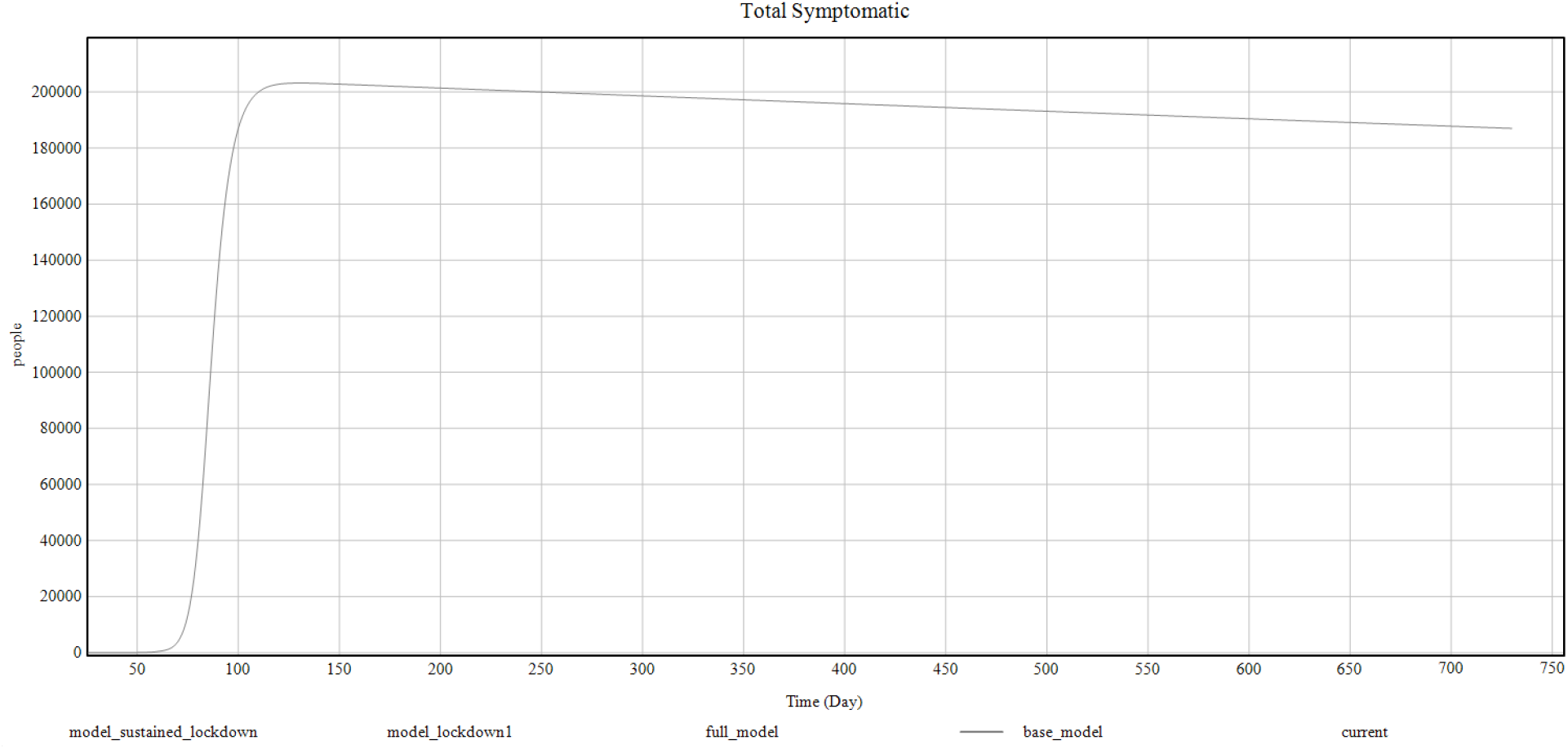
Shows the base model curve for total symptomatic cases against the simulation time period.

### Experiment 2 (Brief Lockdown)

The second experiment assumes the implementation of the 1st set of lockdown measures. It mimics the timings on which the first lockdown was started around day 60 and stopped after 52 days simulation time. This model represents the scenario when only a single down is put in place only to lift it later as happened in Pakistan from March 24 to May 9, 2020 (Table 4).

This model mimics the initial rise of cases in Pakistan closely peaking at around 100000 cases on day 150 and then plateauing subsequently for some time. This model shows a better case then the base model but it predicts a subsequent rise from day 175 if no further lockdown measures are put in place.

**Figure 6.**
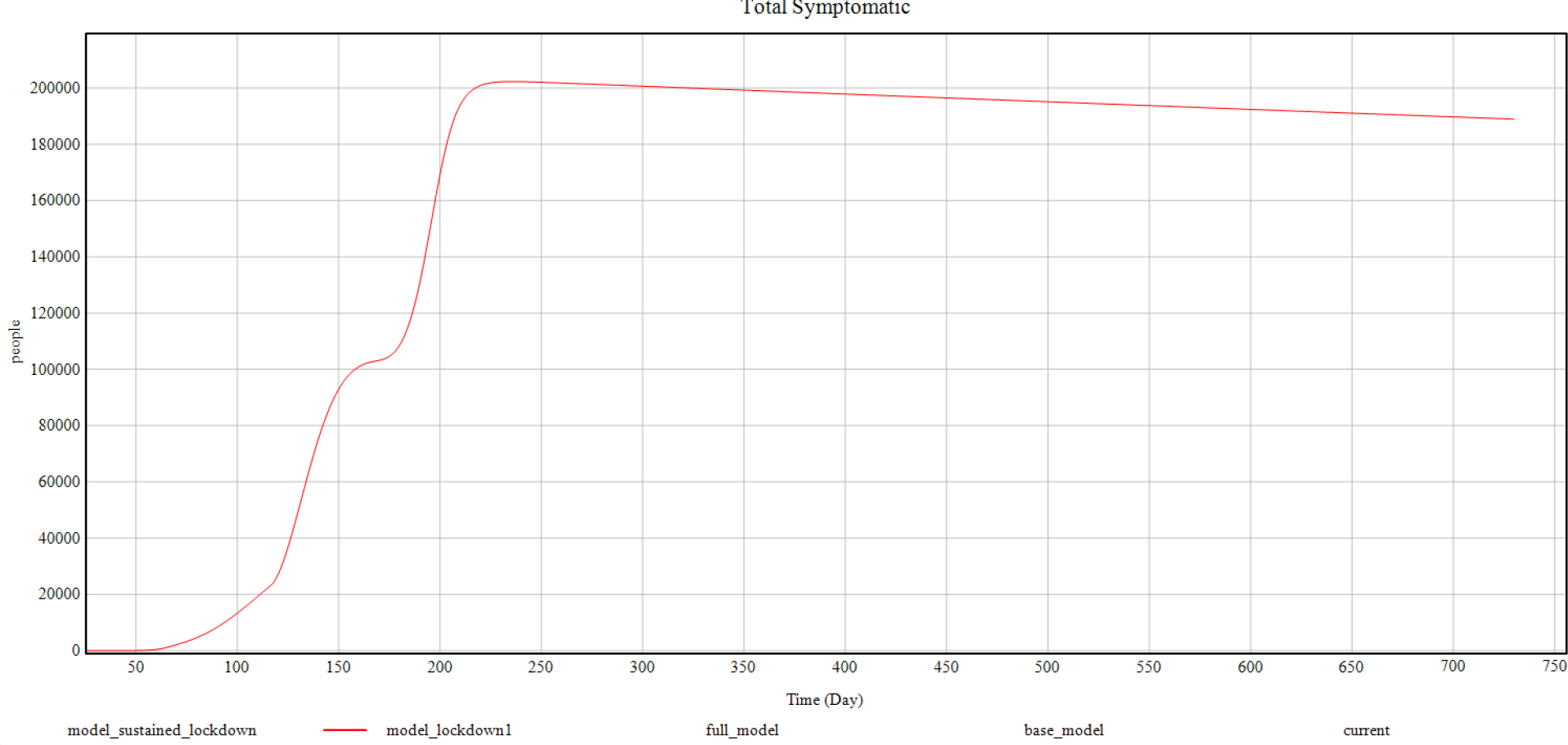
Shows the experiment 2 curve for total symptomatic cases against the simulation time period

### Experiment 3 (Smart Lockdowns)

This model represents the implementation of the second set of lockdown measures also known as the “smart lockdowns”. Since in our model, we assume the population to exist in a single geographic area homogeneously, the concept of smart lockdowns is implemented as the one before. It is evident that the cases follow a similar curve until day 175 but then after a brief plateau, they start to decrease as happened in Pakistan (Table 4).

Since this model assumes the second set of lockdown measures to start around day 162 and persist up to 38 days, the cases start to rise again around day 220 after reaching the lowest case report number of 2829 per day. As per this model, if the smart lockdowns are completely lifted, the cases will rise again and eventually reach a maximum of 200000 cases per day around day 250 simulation time (September 27, 2020).

**Figure 7.**
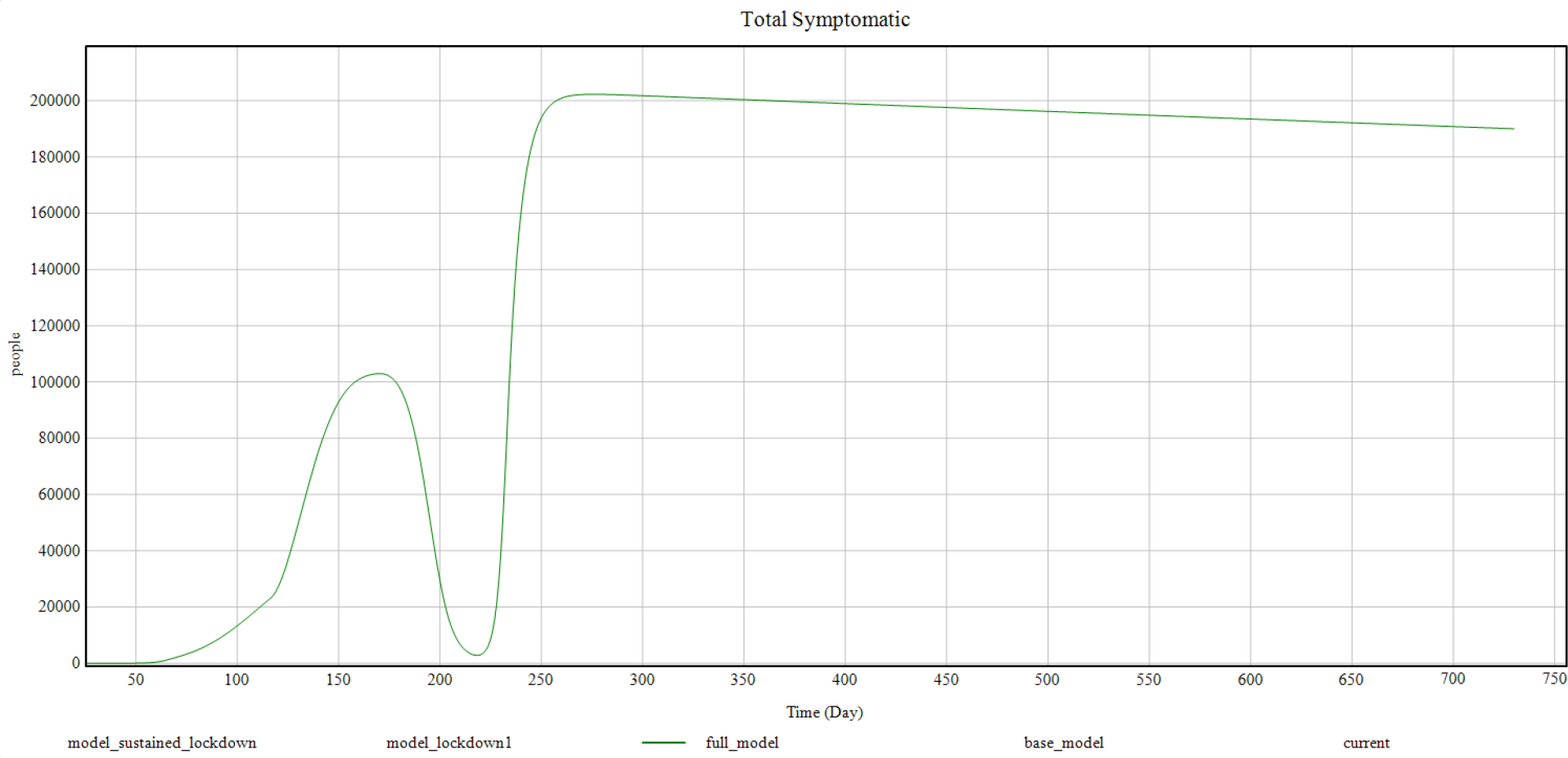
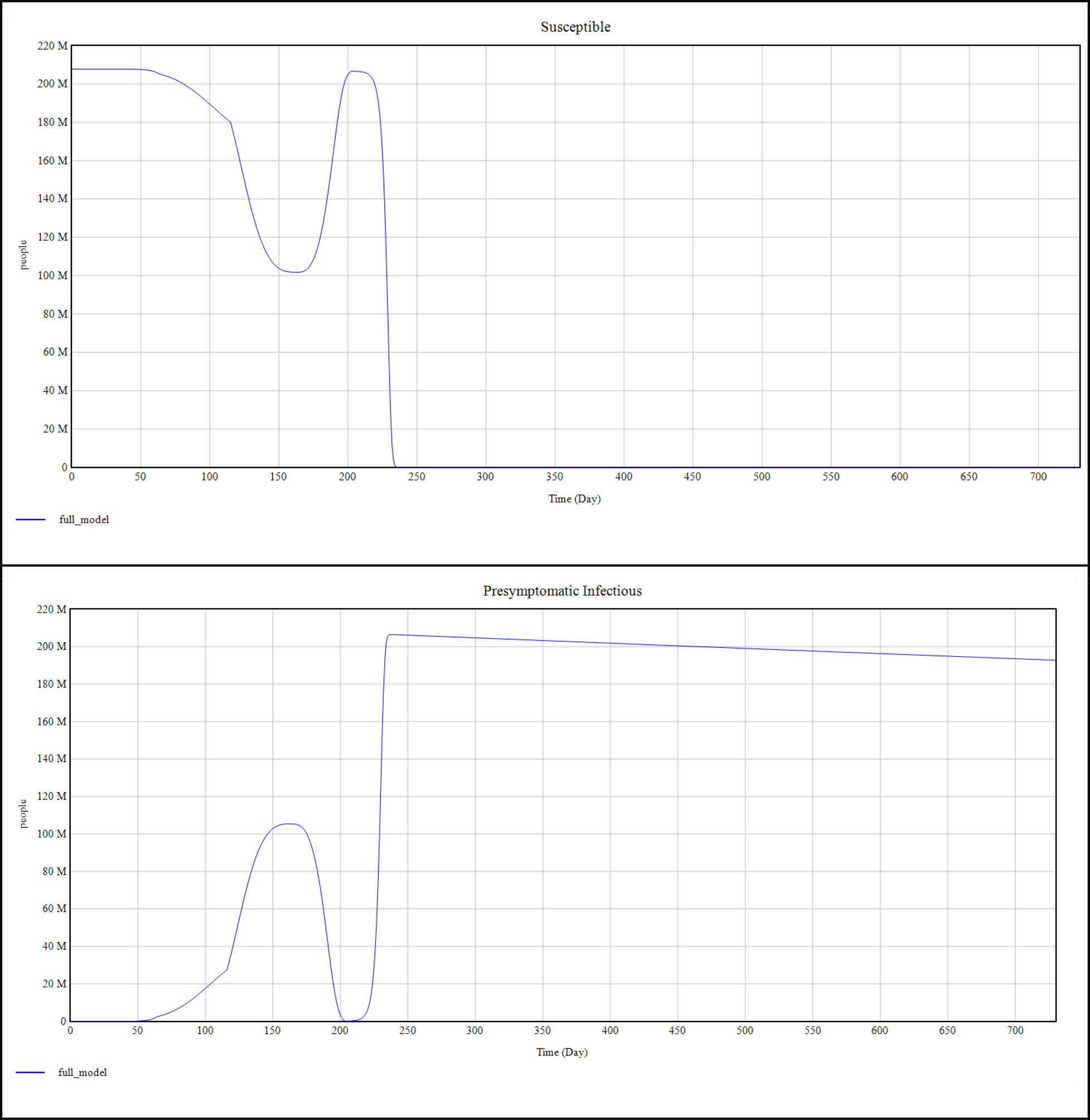
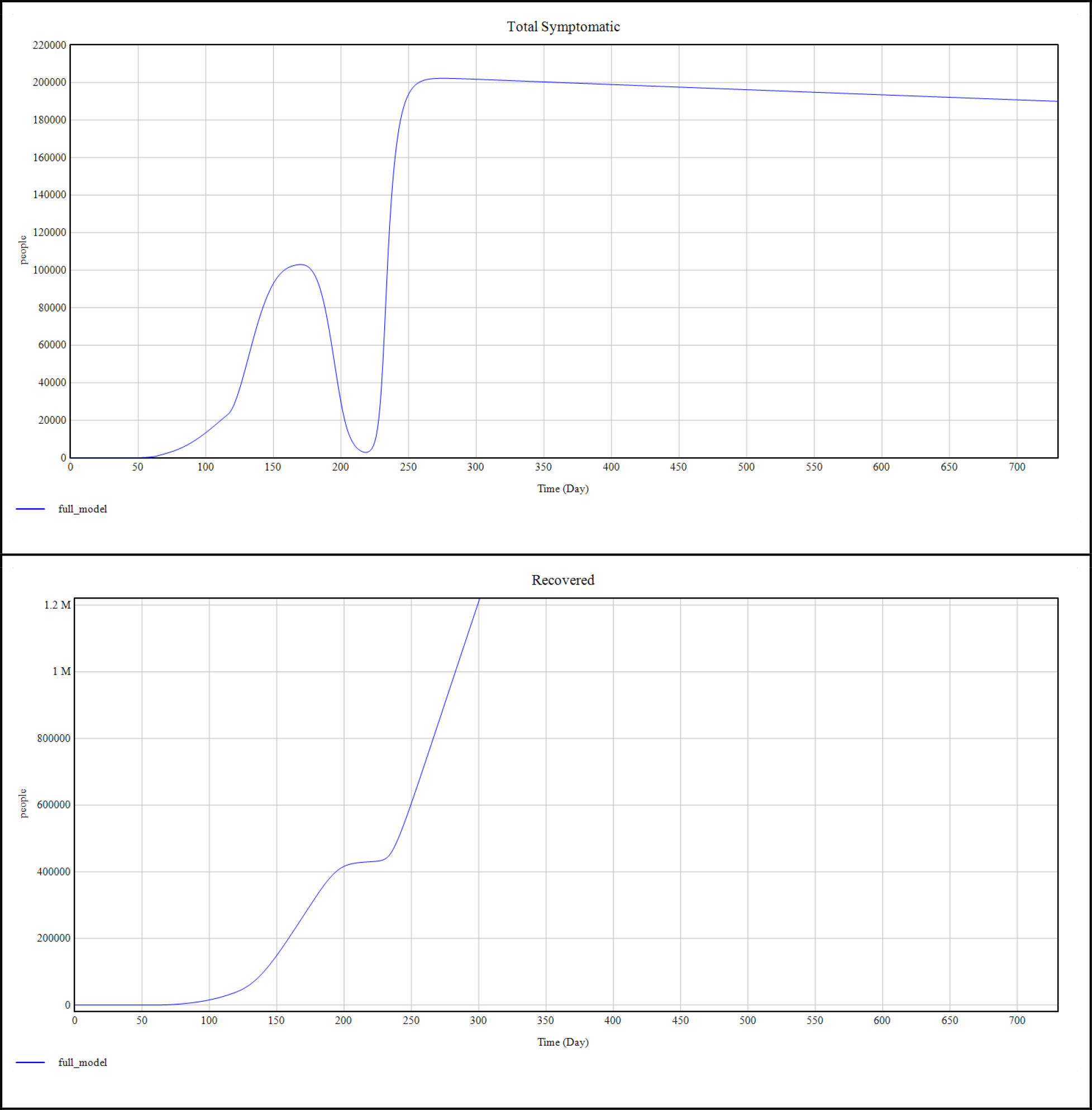
Shows the experiment 3 curve for total symptomatic cases against the simulation time period.

Here we also present the other compartmental graphs as below:

**Figure 8.**
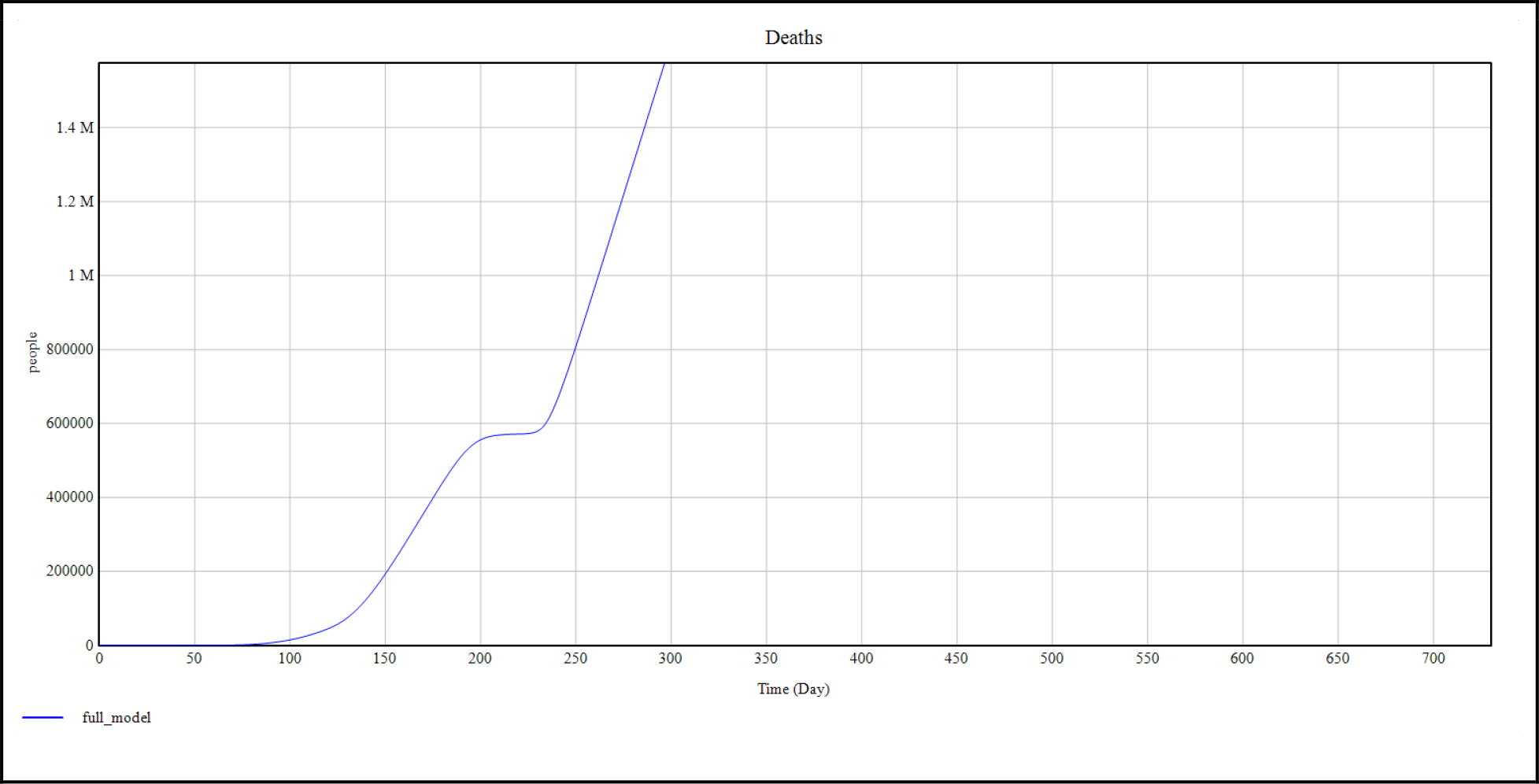
Shows the experiment 3 curves for susceptible, presymptomatic infectious, Total symptomatic, Recovered and Deaths against the simulation time period respectively.

**Figure 9.**
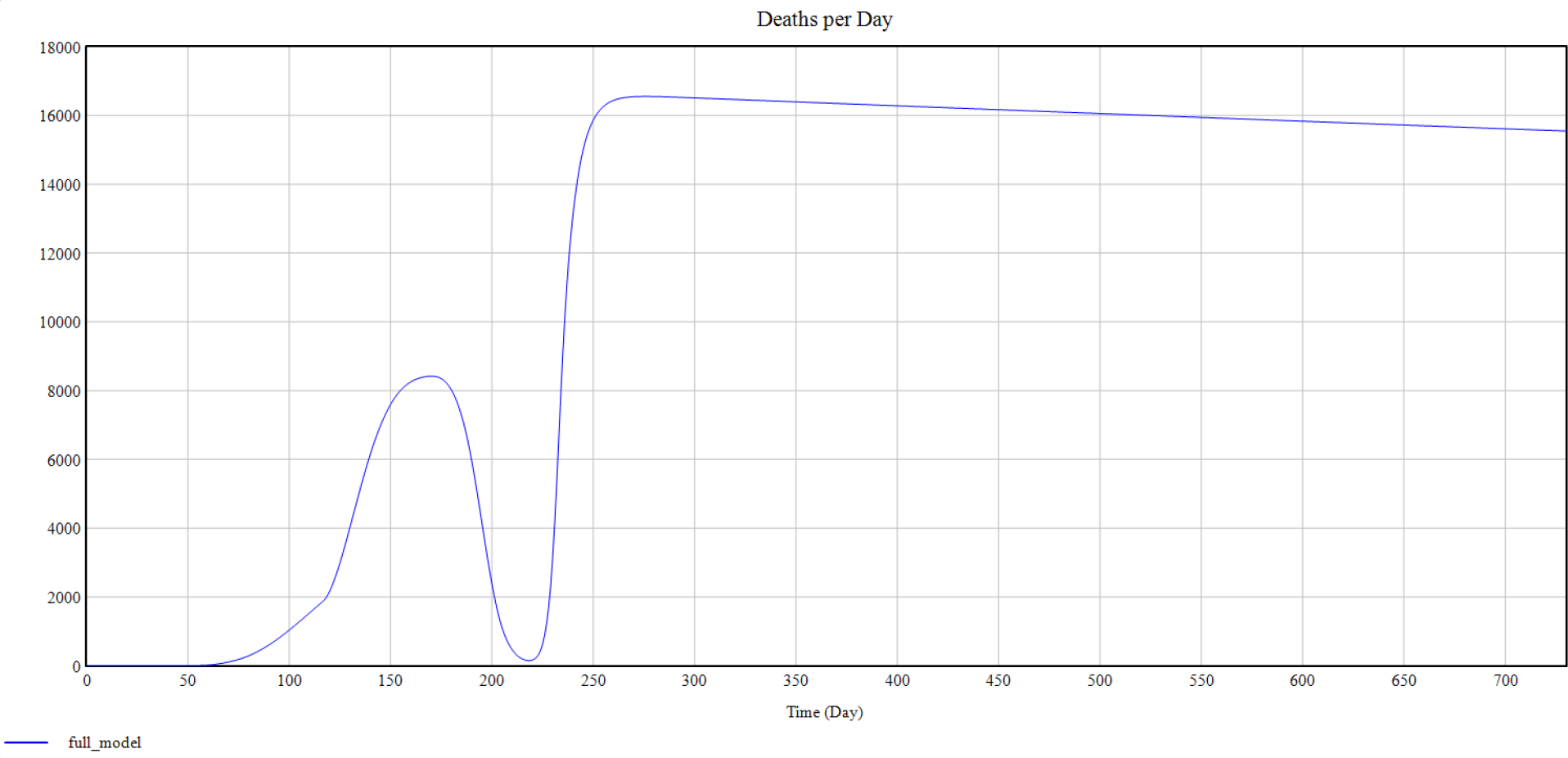
Shows the experiment 3 curve for Deaths per day against the simulation time period respectively.

### Experiment 4 (Sustained Smart Lockdowns)

Continuing from above, If the smart lockdown measures are kept in place with maximum effect, the total number of active cases are shown to drop to less than 1 by day 273 (October 20th, 2020). This model assumes that the lockdown measures don’t lose their efficacy and are kept in place indefinitely which can be difficult in a real world scenario. Any compromise in efficacy of sustained lockdown measures will result in a curve which is intermediate between experiment 3 and 4.

**Figure 10.**
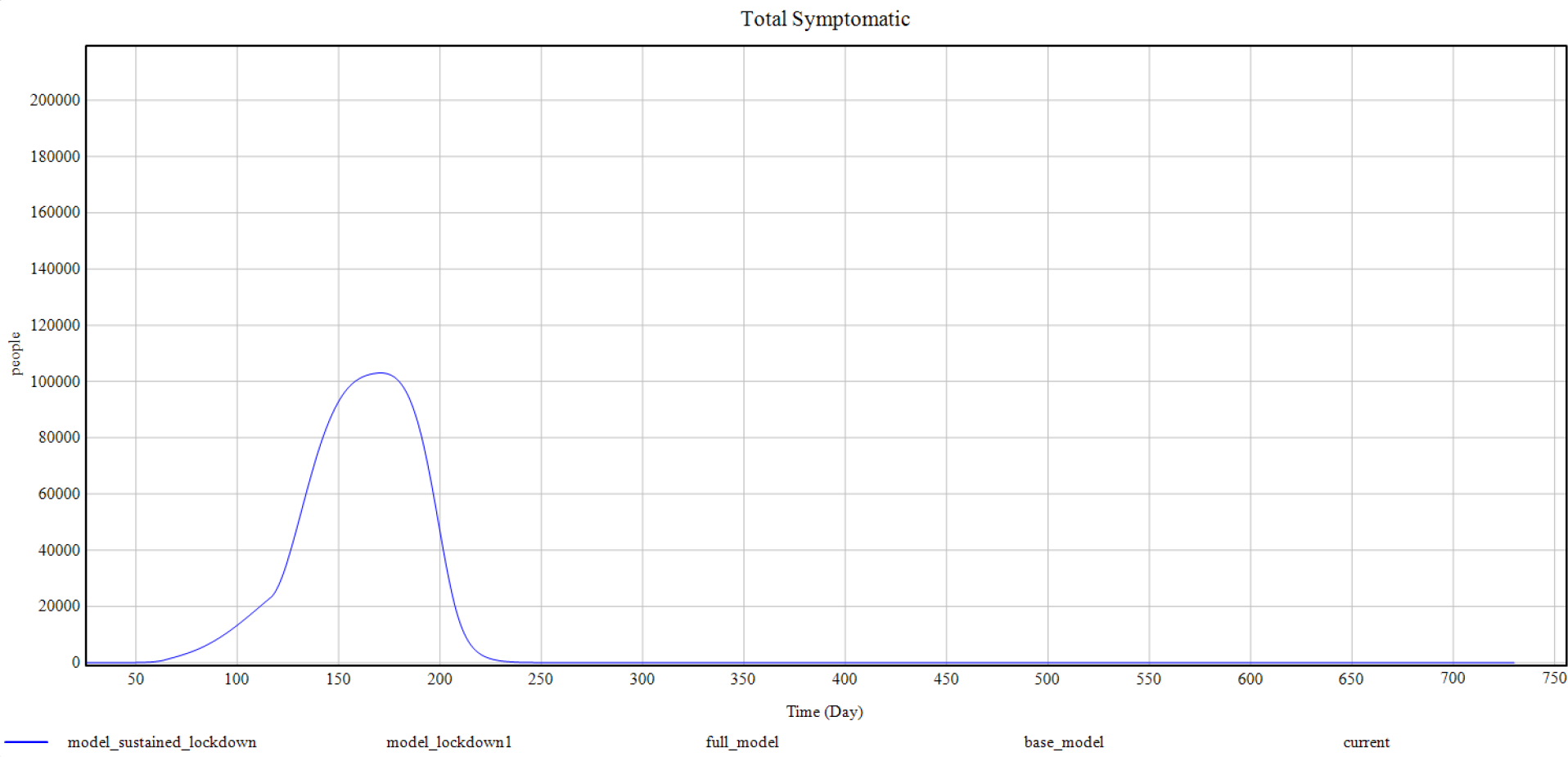
Shows the experiment 4 curve for Total symptomatic cases against the simulation time period

### Experiment 5

Finally, we take the model from experiment 3, assuming that a sustainable option of smart lockdowns is impossible and implement some of the possible solutions into the future.

1. We observe the change in mortality per day by increasing the total number of available ITU beds from an available 4000 (Table 1) to 40000. As in the graph below, the observed change is not much even though there is a definite decrease in mortality because of better respiratory support.
2. As part of a second predictive analysis, we increase the number of available public health beds (includes all available hospital beds and social care services). We increase it from an available number of 124700 to 1 million (Table 1). As shown in the graph below no significant change in total symptomatic cases was observed.
3. As part of final analysis, we introduce a vaccination schedule of 100000 vaccines per day starting from day 220 simulation time (when a rise in cases is expected). As shown in the graphs below, there is an evident decline in the number of active cases per day and in deaths per day though the rate of decline is slow. It shows that without any complementary social distancing / behavioral risk reduction measures, mere act of vaccination would take years for the disease to come under control if the vaccination starts after the second peak is achieved.

**Figure 11.**
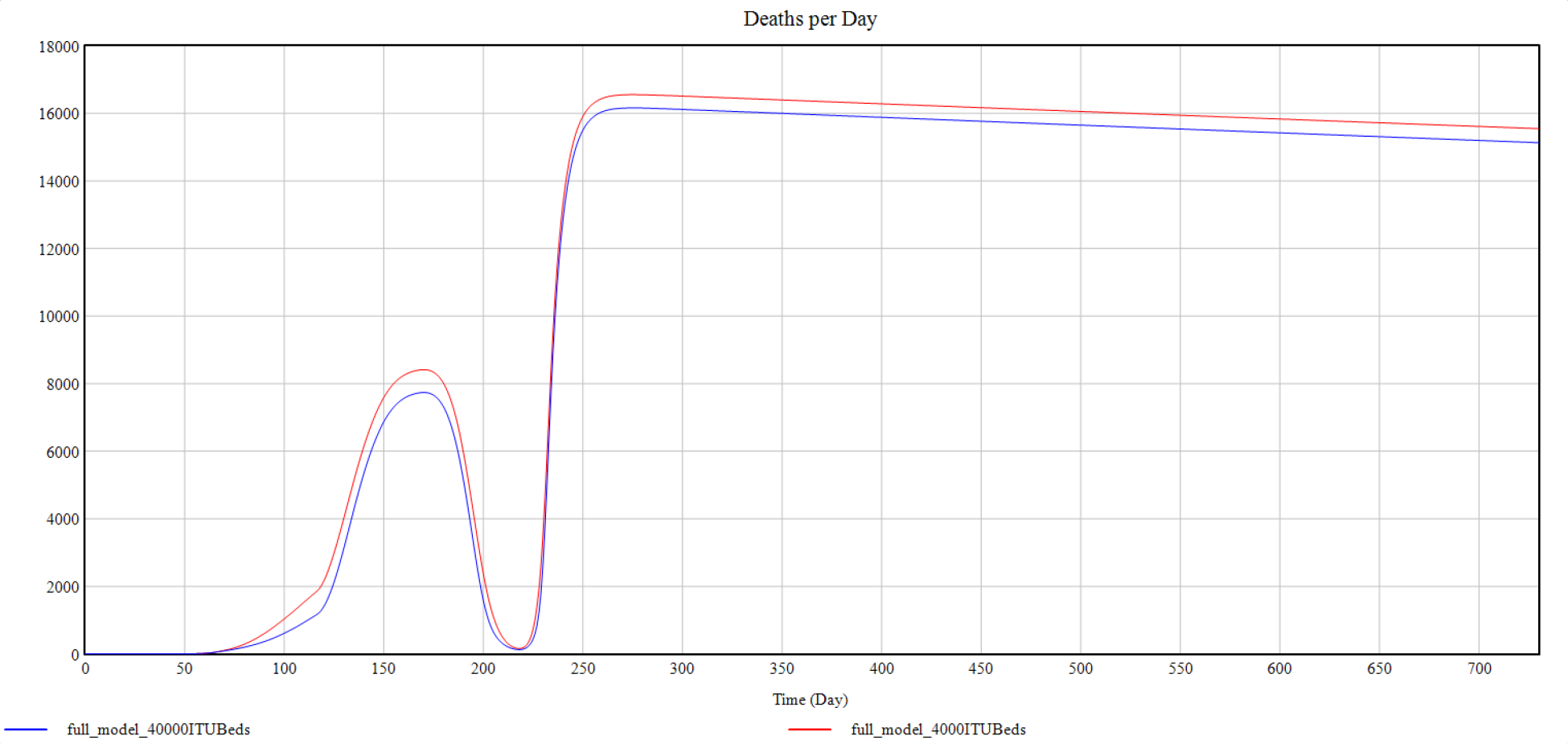
Shows the experiment 5 curve for Deaths per day against the simulation time period comparing scenarios with 4000 vs 40000 ICU bed availability in Pakistan. It shows no significant difference between both scenarios.

**Figure 12.**
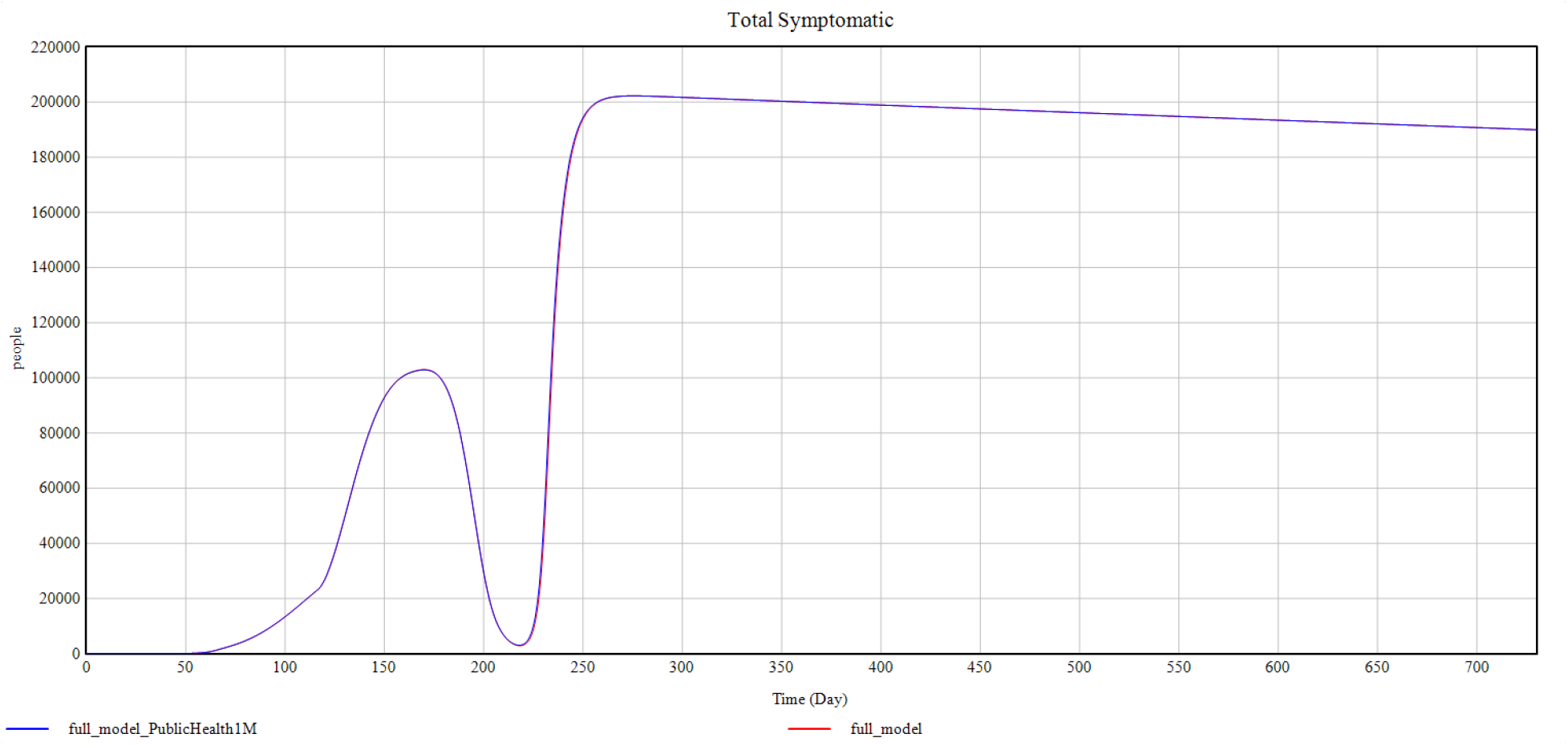
Shows the experiment 5 curve for Total symptomatic cases against the simulation time period comparing scenarios with 124700 vs 1 Million public health capacity units / beds availability in Pakistan. It shows no difference between both scenarios.

**Figure 13.**
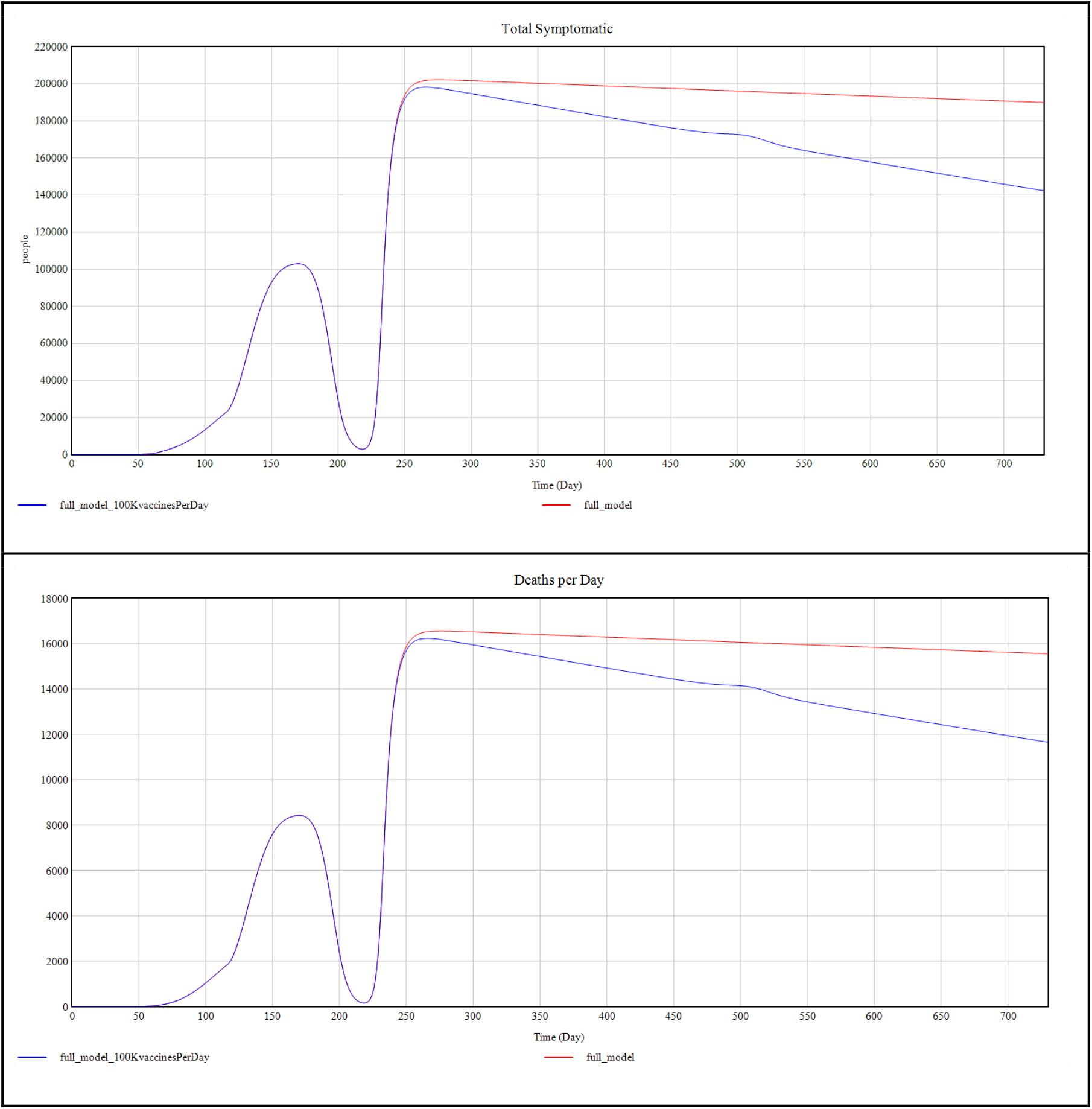
Shows the experiment 5 curves for Total symptomatic cases and Deaths per day against the simulation time period comparing scenarios with none vs 100000 vaccinations per day starting from day 220 simulation time. It shows a visible decline in the incidence of new cases in both curves after the start of vaccination schedule.

## 4. Conclusion

Our study shows the implementation of the SEIDRD model for predicting the incidence of active Covid-19 cases in Pakistan. It tests several experimental scenarios with or without conventional or smart lockdowns, with improvement in bed capacity and with vaccination. It was shown based on the SEIDRD model that after the end of smart lockdowns in Pakistan, the Covid-19 active cases in Pakistan are expected show a rise at the start of September 2020 and if no risk reduction measures are taken, they are expected to achieve a second peak around 27th September 2020. We also observed that increasing the ITU bed capacity to a 10 time current value will not have a significant impact on the number of active cases or mortalities per day. Finally, a vaccination schedule of 100000 vaccines per day started after the second peak of Covid-19 will cause a drop in active cases and mortality per day but the effect will be observed over a period of few years without any risk reduction measures. Based on above findings, we recommend to put in place minimum risk reduction measures (smart lockdowns in high risk areas) at least until the availability of any form of vaccination.

## Data Availability

Real world COVID-19 Data utilized for this study can be found on the following repository maintained by the Center for Systems Science and Engineering (CSSE) at Johns Hopkins University and 'Our World in Data' website page 'Coronavirus Pandemic (COVID-19)' https://github.com/CSSEGISandData/COVID-19
https://ourworldindata.org/coronavirus

## Notes

### Competing Interest Statement

The authors have declared no competing interest.

### Funding Statement

No funding was available provided for this project and was done purely on volunteer basis by the authors

### Author Declarations

Since The research does not contain patient personal data and only publicly available information was used, no IRB / REC approval was sought.

